# Determinants of COVID-19 vaccine acceptance in Saudi Arabia: a web-based national survey

**DOI:** 10.1101/2020.05.27.20114413

**Authors:** Bijaya Kumar Padhi, Mohammed Al-Mohaithef

## Abstract

**Background:** Vaccine hesitancy is a potential threat to global public health. Since, there is an unprecedented global effort to develop a vaccine against the COVID-19 pandemic, much less is known about its acceptance in the community. Understanding key determinants that influence the preferences and demands of a future vaccine by the community may help to develop strategies for improving the global vaccination program. This study aimed to assess the prevalence of the acceptance of COVID-19 vaccine, and their determinants among people in Saudi Arabia.

**Methods:** A web-based, cross-sectional study was conducted using snowball sampling strategy under a highly restricted environment. A bilingual, self-administered anonymous questionnaire was designed and sent to 1000 study participants through social media platforms and email. Study participants were recruited across the country, including the four major cities (Riyadh, Dammam, Jeddah, and Abha) in Saudi Arabia. Associations between COVID-19 vaccine acceptance and sociodemographic profile of the respondents were explored using the chi-squared test. Key determinants that predict vaccine acceptance among respondents were modelled using logistic regression analysis.

**Results:** Of the 1000 survey invitees, 992 responded to the survey (response rate, 99.2%). The majority (65.8%) of the study participants were female, 29.53% were in the age group (36–45 years), and 17.9% were non-Saudi. Of the 992 respondents, 642 (64.72%) showed interest to accept the COVID-19 vaccine if it is available. Willingness to accept the future COVID-19 vaccine is relatively high among older age groups (79.2% among 45+ year old), being married (69.3%), participants with education level postgraduate degree or higher (68.8%), non-Saudi (69.1%), employed in government sector (68.9%). In multivariate model, respondents who were above 45 years (aOR: 2.15; 95% CI: 1.08-3.21), and married (aOR: 1.79; 95% CI: 1.28-2.50) were significantly associated with vaccine acceptance (p <.05). Besides, people having trust in the health system were most likely to accept the vaccine (aOR: 3.05; 95% CI: 1.13-4.92), and those having a higher perceived risk of acquiring infection were 2.13 times (95% CI: 1.35-3.85) higher odds of accepting the vaccine.

**Conclusion:** Addressing sociodemographic determinants relating to the COVID-19 vaccination may help to increase uptake of the global vaccination program to tackle future pandemics. Targeted health education interventions are needed to increase the uptake of the future COVID-19 vaccine.

## Introduction

The severe acute respiratory syndrome coronavirus 2 (SARS-CoV-2) pandemic, which widely referred to as ‘COVID-19’, has been infecting more than 5.5 million over 144 countries(1,2). The pandemic poses a significant threat to the public health system, including catastrophic economic consequences around the world(3,4). Saudi Arabia has been plagued with several pandemics, including the Middle East Respiratory Syndrome Coronavirus (MERS-CoV), and the ongoing COVID-19 outbreak (5,6). As of 26^th^ May 2020, the virus has rapidly spread in the Kingdom, causing a total of 76,726 laboratory-confirmed cases with 411 deaths (CFR 0.53%)(7). A vaccine is considered to be the most awaiting intervention and hundreds of global R&D institutions engaged in unprecedented speed to develop the vaccine (8-11). However, public perception towards COVID-19 vaccine uptake is not available. Numerous studies have shown several factors responsible for vaccine acceptancy when a new vaccine is introduced(12-15). These include the safety and efficacy of the vaccine, adverse health outcomes, misconceptions about the need for vaccination, lack of trust in the health system, lack of knowledge among the community on vaccine-preventable diseases(15,16). Misinformation leading towards vaccine hesitancy could put public health at risk in responding to the current crisis.

In the previous pandemic like the H1N1 influenza A, when the vaccine was introduced, the acceptancy rate varied between 8% to 67% (12). In the United States, the acceptance rate was reported to be 64%(13). In the United Kingdom, 56.1% of the study participants reported accepting the swine flu (influenza A H1N1v) vaccine(17). In Hong Kong, 50.5% of the study population intended to receive a future A/H7N9 vaccine during the outbreak in 2014(18). In Beijing, China, 59.5% of the study participants who had heard of H7N9 were willing to accept a future influenza A(H7N9) Vaccine(19).

Vaccine acceptance and demand are complex in nature and context-specific, varying across time, place, and perceived behavioral nature of the community(12,14,15,20-23). A study in Ireland showed that health care workers avoided seasonal influenza vaccination due to their misconception, efficacy, and trust in the vaccine(24). In China, demographics and public perceptions are the predictors of vaccination acceptance(19). In Hong Kong, anxiety level and vaccine history were the main predictors towards vaccine acceptancy(18). In the United States, perceived effectiveness of the vaccine, social influence, and health insurance was the key predictor towards acceptance of an influenza vaccine(25). Another study in the United States reported greater hesitancy associated with lower vaccine uptake and greater confidence associated with higher vaccine uptake(26). In the United Arab Emirates, a study investigates parent attitudes about childhood vaccines and reported only 12% of parents hesitancy towards childhood vaccination(27). The study reported vaccine safety (17%), side effects (35%), and too many injections (28%) are critical factors in vaccine hesitancy(27). Respondents who had a history of being vaccinated against seasonal flu were more likely to report their intention to be vaccinated(22,23).

A systematic review highlighted the role of public trust in vaccine uptake and reported a dearth in the research of vaccine uptake based on public trust in low and middle-income settings(21). Another review that investigated the general public’s willingness to accept or decline a pandemic vaccine (H1N1) identified several key predictors like people’s perceived risk of infection, the severity of the event, personal consequences, history of previous vaccination, and ethnicity(12). A recent study highlighted that equitable vaccination across all population groups is challenging due to the complex human behavior which changes over space and time (20), and a meta-analysis demonstrated behavioral health model like the “theory of planned behavior” in explaining vaccine hesitancy(14). Numerous studies urged to enhance tailored interventions and policies to increase vaccination uptake(12,14,15,20,24,28).

Few studies has explored the prevalence of COVID-19 vaccine acceptance, and their determinants(29,30). A study conducted among health care workers (HCWs) in China showed a high acceptance of COVID-19 vaccination among health care workers in comparison to the general population(30). Another study in the United States, reported that only 20% intend to decline the COVID-19 vaccine(29). Since vaccine acceptancy is context-specific and varies with geography, culture, and sociodemographic, we aimed to understand the public willingness of a future COVID-19 vaccine in Saudi Arabia.

## Methods

### Study design, sample, and setting

The cross-sectional survey was designed using SurveyMonkey® platform and used a snowball sampling strategy. Study participants were recruited across the Kingdom of Saudi Arabia, including major cities (Riyadh, Dammam, Jeddah, and Abha), and other minor cities. Initially, the study investigators shared the survey link in social media (Twitter, WhatsApp, Telegram channel) and through emails to their primary contacts (aged 18 and above). The primary participants were requested to roll out the survey further. On receiving and clicking the link, participants got auto directed to the informed consent page. After they consented, they were allowed to take the survey. The survey was stopped when it reaches 1000 invitees.

### Study questionnaires

The survey questionnaire was developed in bilingual (Arabic and English) format and consisting sections on sociodemographic, knowledge and perception towards COVID-19, trust in the health system, and participants willingness to accept the COVID-19 vaccine if it is available. The questionnaire was self-administered, and responses were recorded on a Likert scale.

### Data analysis

Descriptive statistics were conducted to generate summary tables for study variables. A cross-tabulation analysis was performed to examine the distribution of intention to uptake COVID-19 with respondents’ sociodemographic characteristics using chi-squared tests. Logistic regression models were employed using a priori hypothesis to tabulate odds ratios (OR) and their 95% confidence intervals (95% CI). All data analysis was performed using STATA 13.0. A two-tailed p-value <0.05 was considered statistically significant.

### Ethical considerations

The study approval was obtained from the Institutional Research Committee of Saudi Electronic University. Study participants were asked to participate voluntarily and were allowed to skip any question if they were not comfortable to answer. Anonymized data was used for analysis and interpretation.

## Results

Of 1000 survey invitees, 992 (99.2%) provided the informed consent and returned the survey. Table 1 shows the summary statistics of the sociodemographic profile of the study participants. Most of the respondents 436 (43.9%) were aged between 26-35 years, followed by 264 (26.6%) aged 18-25 years, 239 (24%) aged 36-45 years, and 53 (5%) were aged 45 years and above (Table 1). Of the 992 participants, 653 (65.8%) were female, 512 (51.6%) were married, majority 814 (82%) of the study participants were Saudi nationality, 455 (45.8%) were from Riyadh, 497 (50.1%) completed graduate-level education, and 428 (43.1%) reported working in a government sector (Table 1).

**Table 1:**
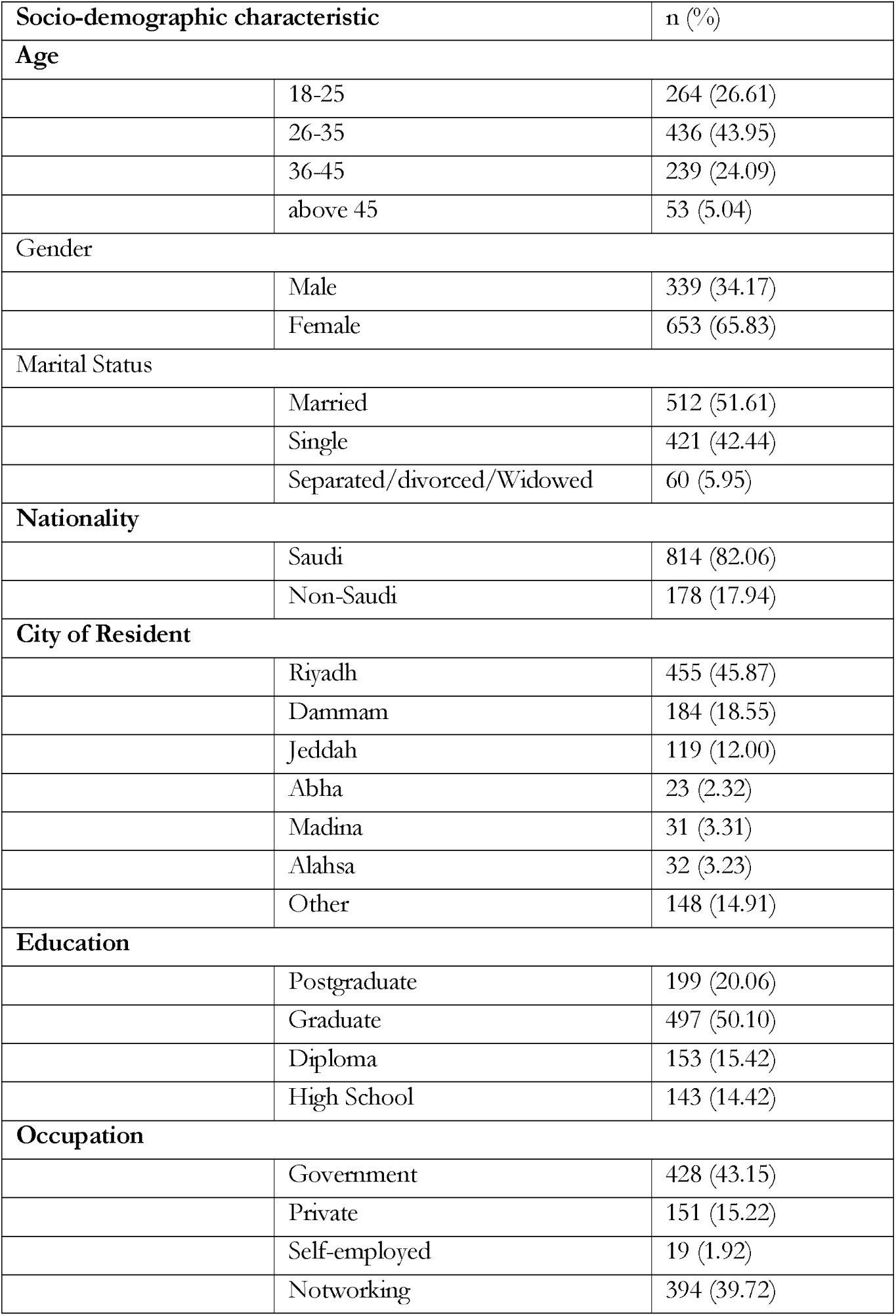
Socio-demographic characteristics of the study population (N=992)

Table 2 shows bivariate associations between sociodemographic characteristics and intent to uptake the COVID-19 vaccine among respondents in Saudi Arabia. Of the 992 respondents, 642 (64.7%) intended to uptake the hypothetical vaccine, only 70 (7%) reported hesitancy towards the COVID-19 vaccine, and 280 (28.2%) were reported “not sure” about their intention (Table 2). Of the 53 respondents who were aged 45 years and above, 42 (79.2%) of them showed interest to uptake the vaccine if it is available. Of the 512 participants who were married, 355 (69.3%) reported accepting the COVID-19 vaccination (Table 2).

**Table 2:** Bivariate associations between socio-demographic characteristics and intent to uptake the COVID-19 vaccine among respondents in Saudi Arabia (N=992)

| Variables | “If Vaccine against Coronavirus is available, I will take it” |  |  |  |
| --- | --- | --- | --- | --- |
|  | Yes<br>(n=642) | No<br>(n=70) | Not sure<br>(n=280) | p-value |
| **Age** |  |  |  |  |
| 18-25 | 172 (65.15) | 14 (5.30) | 78 (29.55) | 0.201 |
| 26-35 | 280 (64.22) | 36 (8.26) | 120 (27.52) |  |
| 36-45 | 148 (61.92) | 19 (7.95) | 72 (30.13) |  |
| above 45 | 42 (79.25) | 1 (1.89) | 10 (18.87) |  |
| **Gender** |  |  |  |  |
| Male | 235 (69.32) | 22 (6.49) | 82 (24.19) | 0.087 |
| Female | 407 (62.33) | 48 (7.35) | 198 (30.32) |  |
| **Marital status** |  |  |  |  |
| Single | 248 (58.91) | 33 (7.84) | 140 (33.25) | 0.024 |
| Divorced/Widowed | 39 (66.10) | 4 (6.78) | 16 (27.12) |  |
| Married | 355 (69.34) | 33 (6.45) | 124 (24.22) |  |
| **Nationality** |  |  |  |  |
| Saudi | 519 (63.76) | 63 (7.74) | 232 (28.50) | 0.152 |
| Non-Saudi | 123 (69.10) | 7 (3.93) | 48 (26.97) |  |
| **City of residence** |  |  |  |  |
| Riyadh | 286 (62.86) | 36 (7.91) | 133 (29.23) | 0.323 |
| Dammam | 113 (61.41) | 10 (5.43) | 61 (33.15) |  |
| Jeddah | 80 (67.23) | 7 (5.88) | 32 (26.89) |  |
| Others | 163 (69.66) | 17 (7.26) | 54 (23.08) |  |
| **Highest education** |  |  |  |  |
| High School | 88 (61.54) | 10 (6.99) | 45 (31.47) | 0.472 |
| Diploma | 99 (64.71) | 7 (4.58) | 47 (30.72) |  |
| Under Graduate | 318 (63.98) | 42 (8.45) | 137 (27.57) |  |
| Post Graduate | 137 (68.84) | 11 (5.53) | 51 (25.63) |  |
| **Current occupation** |  |  |  |  |
| Not working | 242 (61.42) | 29 (7.36) | 123 (31.22) | 0.145 |
| Private/Self-employed | 105 (61.76) | 11 (6.47) | 54 (31.76) |  |
| Government | 295 (68.93) | 30 (7.01) | 103 (24.07) |  |

Table 3 presents logistic regression analysis for sociodemographic prediction of intent to uptake the COVID-19 vaccine among respondents. In the multivariate model, respondents who were above 45 years are 2.15 times likely to accept the vaccine (aOR: 2.15; 95% CI: 1.08-3.21). Similarly, participants who were married are 1.79 times likely to accept the vaccination (aOR: 1.79; 95% CI: 1.28-2.50).

**Table 3:**
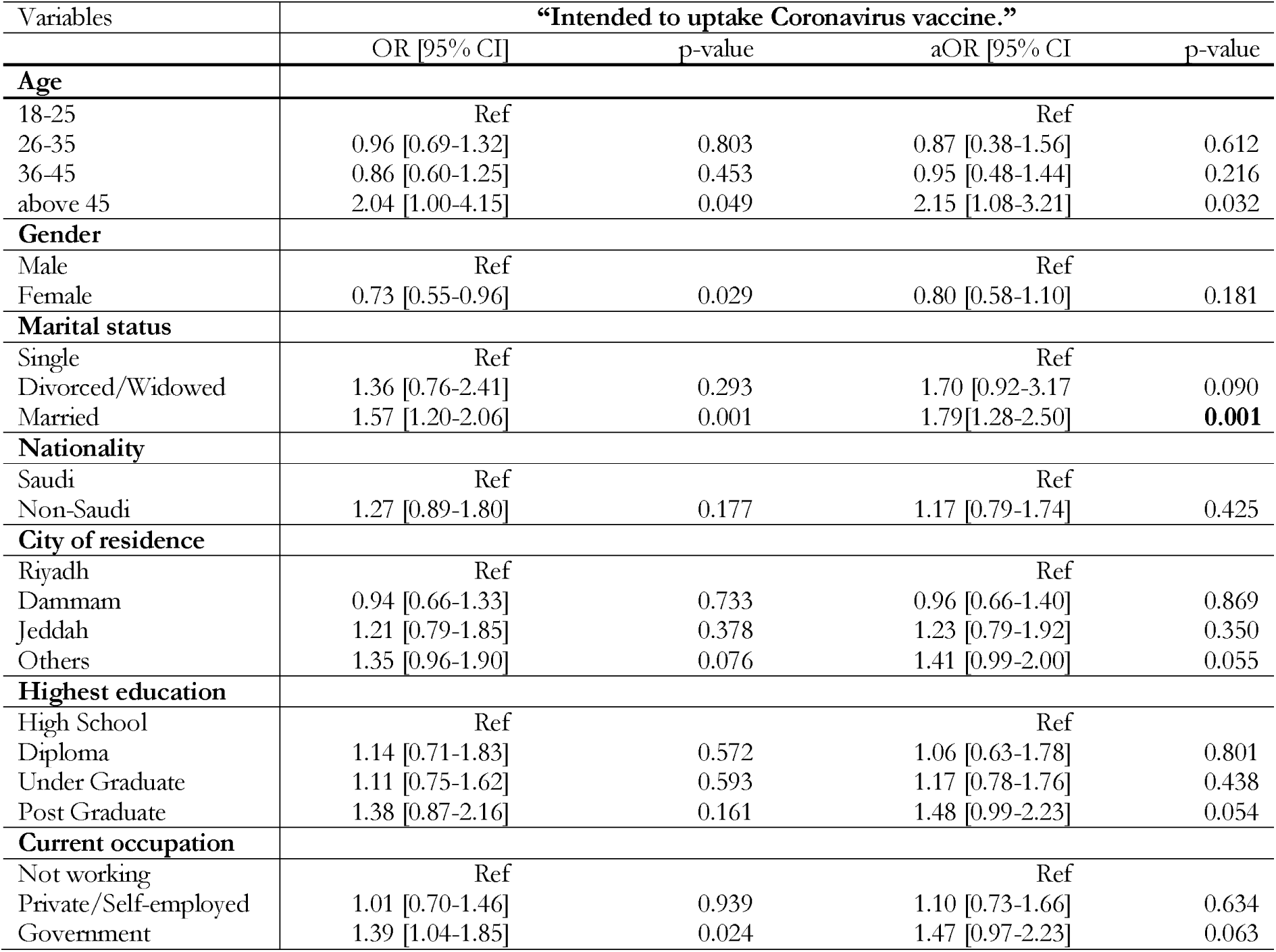
Logistic Regression analysis for socio-demographic prediction of intent to uptake the COVID-19 vaccine among respondents in Saudi Arabia (N=992)

Table 4 shows the logistic regression analysis for factors potentially associated with the intention to receive the COVID-19 vaccine among respondents. In the multivariate model adjusted for sociodemographic characteristics, participants who concerned about acquiring infection with the COVID-19 virus were 2.13 (95%CI: 1.35-3.85) times likely to accept the COVID-19 vaccine compared with those who were not concerned with the infection. Participants, who trusted the health system were 3.05 (95%CI: 1.13-4.92) times most likely to accept the vaccination than those who have reported no trust.

**Table 4:** Logistic Regression analysis for factors potentially associated with the intention to receive COVID-19 vaccine among respondents in Saudi Arabia (N=992)

| Variables | “Intended to uptake Coronavirus vaccine.” |  |  |  |
| --- | --- | --- | --- | --- |
|  | OR [95% CI] | p-value | aOR [95% CI] | p-value |
| **Perceived risk** |  |  |  |  |
| No | Ref |  | Ref |  |
| Yes | 2.48 [1.11-3.95] | 0.031 | 2.13 [1.35-3.85] | 0.002 |
| **Trust in the health system** |  |  |  |  |
| No | Ref |  | Ref |  |
| Yes | 2.85 [1.03-4.80] | 0.028 | 3.05 [1.13-4.92] | 0.001 |

## Discussions

Vaccination is considered one of the most outstanding public health inventions in the 21^st^ century. However, its acceptancy is varied with space, time, social class, ethnicity, and contextual human behavior (14,15,20,21,31). Our study, first of its kind in Saudi Arabia, used a web-based self-administered questionnaire and collected responses across the Kingdom, including four major cities (Riyadh, Jeddah, Dammam, and Abha) and some minor cities in the country. Of the 992 study participants, 642 (64.7%) said ‘yes’ to uptake the COVID-19 vaccine, 70 (7.0%) said ‘no’ to uptake COVID-19 vaccine, and 280 (28.2%) said ‘not sure’ to uptake the COVID-19 vaccine if it is available. Further, being aged (45 years and above) (aOR: 2.15; 95% CI: 1.08-3.21), and being married (aOR: 1.79; 95% CI: 1.28-2.50) are likely to accept the COVID-19 vaccine than their counterparts. Study participant’s trust in the health system (aOR: 3.05; 95% CI: 1.13-4.92) and perceived risk of acquiring infection (aOR: 2.13; 95% CI: 1.35-3.85) were found to be significant predictors in explaining acceptancy of the COVID-19 vaccine.

Though there have been limited studies to explore the intention to uptake the COVID-19 vaccine in the current crisis, our results are in agreement with study conducted in China, and in the United States(29,30). The Chinese study reported 72.5% of the general population’s intention to uptake COVID-19 vaccine(30). And the study conducted in the United States reported 80% acceptance of the COVID-19 vaccine among the study population(29). In our study, 64.7% of study participants showed interest in the uptake of the COVID-19 vaccine. Similar observations were made during the H1N1 pandemic(12).

Some qualitative comparisons can be made with similar studies, like in a systematic review, the acceptance rate varied between 8% to 67% for the H1N1 influenza A pandemic vaccine(12). The acceptance rate was reported to be 64% in the United States(13), 56.1% in the United Kingdom (17), 59.5% in Hong Kong(18), and 59.5% in China (19). The systematic review also highlighted that there was no consistent association with participants’ demographic variables (age and sex) with vaccine uptake behavior (12). However, in our study, old aged participants are more likely to accept COVID-19 vaccination than their counterparts (aOR: 2.15; 95% CI: 1.08-3.21).

Numerous studies reported the perceived risk of becoming infected as a predictor towards intention behind vaccination(12,22,23,30). In our study, participants who had a higher perceived risk of being infected are 2.13 times more likely to be vaccinated than those having a lower perceived risk (aOR: 2.13; 95% CI: 1.35-3.85). Studies have shown that a higher trust in the health system is associated with the utilization of preventive health services such as vaccination(26,32,33). In our study, odds of having greater trust in the health system were 3.05 times higher, reporting their intention to uptake the COVID-19 vaccine (aOR: 3.05; 95% CI: 1.13-4.92).

Our study has several limitations; firstly, it is cross-sectional, depicts a picture of the community response at the point of the study. We asked the respondents to report their intention to receive the COVID-19 vaccine if it is available in the future. A considerable number of study participants (28.2%) reported “Not sure” about their intention to uptake the COVID-19 vaccination. The real intention could be different when the vaccine is available(22). It is interesting to study how the intention varies over time and the context in the study population. Secondly, study responses were recorded using a web-based self-administered survey, instead of a direct face-to-face interview. This may lead to potential bias in reporting their responses. Third, the current study didn’t explore the motivation behind the acceptance or barriers behind the hesitancy of the VOVID-19 vaccine.

Despite the above limitations, our study is the first of its kind, with a representative sample size across the county demonstrated the population’s intention to uptake the COVID-19 vaccine. Once the pandemic is over, we will explore many additional research questions, including vaccine promotion strategies, vaccine safety, vaccine referral/recommendations, cost (out of pocket expenditure), including the key motivation and barriers towards COVID-19 vaccination.

## Conclusions

This is the first community-based study under a highly restricted environment that assessed the public’s intent to accept the hypothetical COVID-19 vaccine in the Kingdom with a representative sample. The study participant has a good intention to accept the hypothetical vaccine and is in accordance with the previously reported figures. Participants’ perceived risk and trust in the health system were found to be significant predictors towards the intention of the COVID-19 vaccine in the Kingdom. Further study should corroborate our findings with public health promotion interventions.

Health education targeting various sociodemographic groups should be taken as a priority to increase the COVID-19 vaccine uptake behavior in the country, and elsewhere.

## Data Availability

The Data is available with the corresponding author and can be provided upon request.

## Acknowledgments

The authors would like to acknowledge the research support provided by Saudi Electronic University.

## Data Availability Statement

The data that support the findings of this study are available from the corresponding author upon request.

## Author Contributions

Conceived and designed the experiments: MAM, BKP

Performed the experiments: MAM

Analyzed data: BKP, MAM.

Wrote the manuscript: BKP, MAM

Review and editing of the manuscript: MAM, BKP

